# Assessment & mitigation of O2 therapy driven spread of COVID-19

**DOI:** 10.1101/2021.02.06.21251266

**Authors:** Arshad Kudrolli, Brian Chang, Jade Consalvi, Anton Deti, Christopher Frechette, Helen Scoville, Geoffrey R. Sheinfeld, William T. McGee

## Abstract

**BACKGROUND:** Exhalation exposure from patients to healthcare workers (HCWs), while using a nasal cannula or simple O2 mask used in treating COVID-19 and other respiratory diseases, is a present and future risk. Little is known on exhalation dispersal through these devices, and on mitigating the viral exposure to those in the vicinity.

**METHODS:** Respiration through O2 therapy devices was studied with a supine manikin equipped with a controllable mechanical lung by measuring aerosol density and flow with direct imaging. Dispersal direction and distances were quantified while placing a surgical mask loosely over the devices and contrasted with unmitigated oxygenation device use. Exhalation jets were examined over the entire range of oxygenation rates used in treatment.

**RESULTS:** Exhalation jets travel 0.35 ± 0.02 m upward while wearing a nasal cannula, and 0.29 ± 0.02 m laterally while wearing a simple O2 mask posing significant inhalation risk. Placing a surgical facemask loosely over the oxygenation device is demonstrated to alleviate exposure by reducing and deflecting the exhalation jets from being launched forward, and by reducing exhalations from being launched directly higher over a supine patient. Less than 12% of the exhaled breath is observed to reach above a masked face where HCWs would be present, independent of oxygen flow rates.

**CONCLUSIONS:** Exhalation jets from both the nasal cannula or simple O2 mask were found to concentrate aerosol-laden exhalations directly in front of a patient’s face. Exposure is effectively mitigated with a surgical mask which reduces and redirects the exhalation downward away from HCWs.

## 1 Introduction

Health care workers (HCWs) have been known to contract COVID-19 from exposures at work [1, 2, 3]. Sources of exposure include coworkers as well as the patients they care for. It is assumed that surgical face masks suffice for prevention of viral transmission from respiratory droplets, while N95 respirators provide additional protection from airborne transmission via bioaerosols [4, 5]. Evidence for various comparisons about masks used in health care settings and risk for COVID-19 remains insufficient [6].

Oxygen therapy is the major treatment modality for seriously ill patients with COVID-19 [7]. For most hospitalized patients with COVID-19, oxygen therapy is provided by either a nasal cannula or a simple O2 mask (see Supplementary Information A). Exhalation jets emerging from masks with vents are well documented [8] and are not recommended by the CDC in public settings because they do not prevent the spread of infection. Nonetheless, hospitalized COVID-19 patients receiving oxygen by either of these methods rarely use further mitigation to prevent the spread of exhaled aerosols and droplets. A national survey of 37 hospitals with 115 responses found less than 10% use of face masks according to 60% of responses and over 75% rates of face mask use for only 5% of responses for patients known to be infected with COVID-19 and using supplemental oxygen [9]. Aerosol-laden exhalations through oxygenation devices and its impact on infectious disease spread have been studied [10, 11], and informed early strategies used in treating patients by intubation [12]. Poor outcomes following intubation has driven care to less invasive oxygen therapy and protecting clinicians and other caregivers with personal protection equipment (PPE) [13]. Nonetheless, infections to health care workers remains a significant concern [14], and clinicians continue to get infected in large numbers compared with the general population [2].

Aerosol dispersal mitigation by a surgical mask over high flow oxygen therapy has been proposed [15] and preliminary clinical data is available for mitigation of aerosols close to the patient where bedside care is delivered [16, 17]. A clear demonstration of mask efficacy worn by patients with more commonly used oxygen delivery devices remain unclear and is not practiced widely [9]. In this study, we demonstrate that a loosely placed surgical mask over a nasal cannula, or simple O2 mask, decreases and redirects exhalations downward, and thus away from the faces of caregivers, while simultaneously reducing the volume and density of the exhaled aerosol.

## 2 Methods

A novel manikin exhalation system enables us to visualize and quantify the direction and density of aerosol-laden exhalations of patients being treated with oxygen therapy under prescribed and reproducible conditions, which is impossible with actual human subjects. A schematic and image of our experimental apparatus can be found in Supplementary Information A. A Michigan Instruments Dual Lung Simulator and a ventilator (ParaPAC plus 310) configured to mimic negative pressure spontaneous patient respiration with a prescribed tidal volume and frequency was used. A tube connects the Lung Simulator with a Head Simulator Module (HSM-A) via which the manikin breaths through the nose and/or mouth. A tidal volume of *V*_*t*_ = 500 mL with a breathing rate of *f* = 12 breaths per minute (bpm) represents normal breathing. Patients that require oxygen assistance typically have a lower tidal volume which is compensated with a higher frequency [18]. Thus, a tidal volume of *V*_*t*_ = 350 mL and respiratory rate of *f* = 20 bpm simulated a patient with lung disease, and models typical COVID-19 patients who are experiencing shortness of breath while undergoing supplemental oxygen therapy in our practice. Metered O_2_ from a tank is delivered through a nasal cannula (Vyaire) or simple O2 mask (Vyaire) with flow rates *Q* listed in the table in Fig. 1 typically used in treating Covid-19 and other respiratory disease patients. All measurements were conducted in a room with HVAC at 23.5°C with standard deviation of 0.5°C, and humidity 21.0 % with standard deviation of 2.5 %.

**Figure 1:**
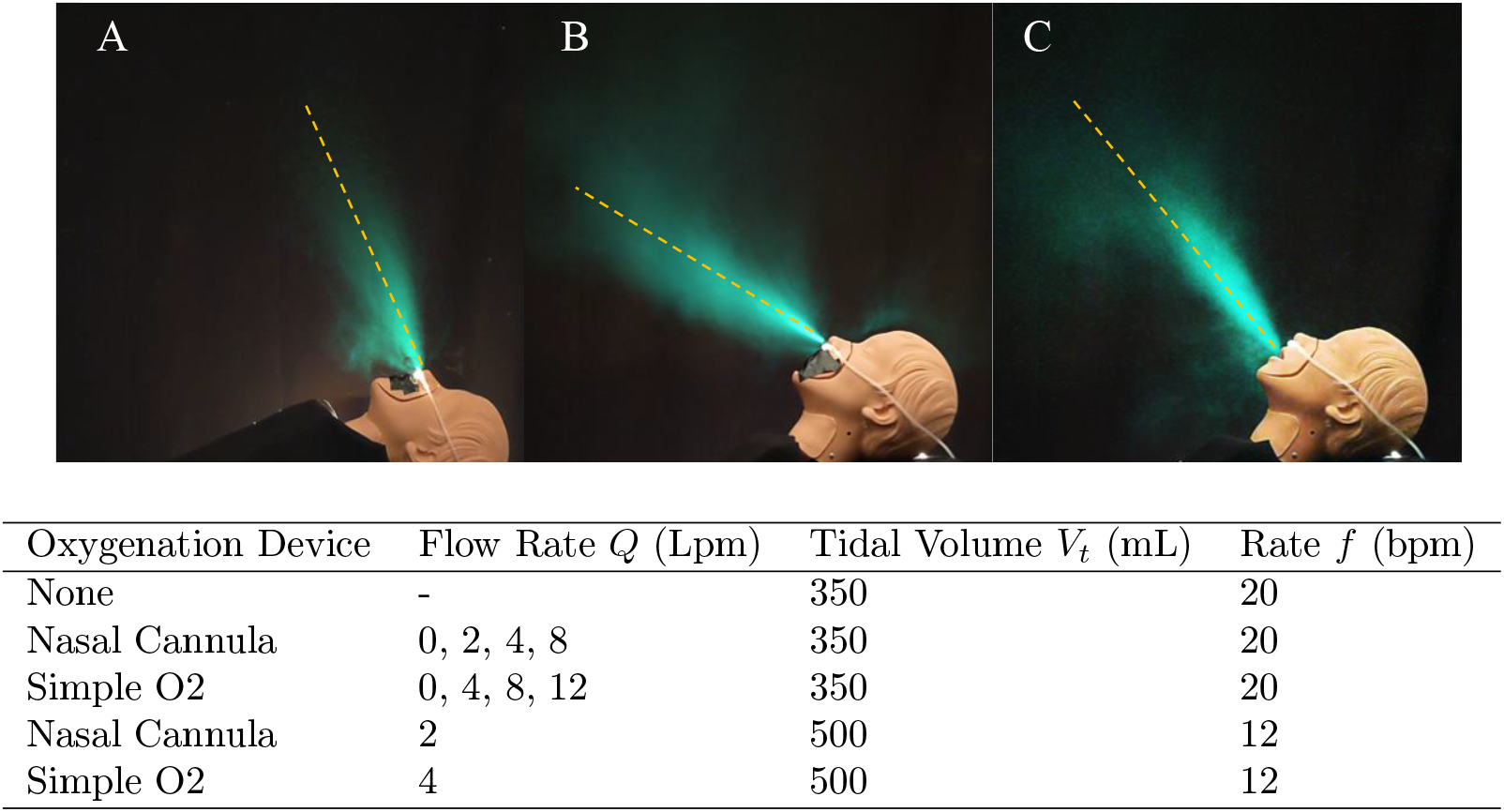
The time averaged exhalations emerging from the nose of a medical manikin fitted with a nasal cannula when the head rests flat (A), and while it is on a bed propped at 45 degrees (B), and when exhaling through the mouth (C), under shortness of breath conditions (*V*_*t*_ = 350 mL at *f* = 20 bpm; *Q* = 4 Lpm). The aerosol-laden exhalation is visualized with back lighting and rendered with artificial cyan coloring. A primary turbulent exhalation jet is observed which spreads conically with principal direction indicated with dashed line with elevation angle *θ* = 60 ± 0.5 degrees (A), 19.3 ± 0.5 degrees (B), and 56 ± 1 degrees (C). Table: Summary of the oxygenation devices, flow rates, and breathing tidal cycles investigated (*Q* = 0 Lpm cases were conducted for calibration purposes). Corresponding movies found in SI Movie 1-3.

In order to image the exhalations, we use an aerosol fog composed of approximately 1-5 micron water-based droplets which scatter light while moving with the air flows [10, 19]. While the droplet sizes are optimized for light scattering and more numerous in number, they can be noted to be within the range of bioaerosols exhaled while breathing [20, 21, 10]. Two complementary lighting methods are used to obtain the overall direction and spread of the aerosol-laden exhalations and in deducing where the flow is jet-like with linear momentum, versus spreading diffusively as they slow down. We place a 5500 lumen LED white light source behind the head to backlight the aerosols in the exhalations [22, 23]. Additionally, we use a green laser sheet (532 nm, 40 mW) to visualize the flow in a 2D plane [24]. A Pixel 4a featuring a 12.2-megapixel camera is used to capture movies with 1080p at 30 frames per second (fps) over several exhalation cycles. All quantitative analysis is performed with at least 5 trials for each set of parameters. The image processing methods used to map the image intensities to exhalation density and flow characteristics can be found in Supplementary Information B. The shape of the exhalation jet from the nose and mouth of the manikin can be observed to be typical as when a fast moving fluid enters and loses momentum in a quiescent-fluid [25, 26]. Tracking the leading edge of an exhalation jet over consecutive frames, we observed nasal exhalation with a speed of 1.21 ± 0.07 m/s, consistent with the 0.4 to 1.6 m/s range reported for normal human nasal breathing [21].

## 3 Results

### 3.1 Dispersal through Nasal Cannula

Figure 1 shows time-averaged exhalations emerging from the manikin while undergoing oxygenation treatment with a nasal cannula under varying conditions over five breath cycles each. The corresponding movies can be found in the supplementary documentation SI Movies 1-3 [27]. In each case a primary jet can be observed clearly extending from the nose past the nasal cannula as it enters and spreads conically in the relatively still-air in the room while losing speed (see also Supplementary Information C). Comparing Fig. 1A and Fig. 1B, we observe that the jet on average emerges from the nose somewhat similarly in relation to its face, no matter its angle of tilt, i.e., the direction of the exhalation jet is essentially set by the direction of the face. While wearing a nasal cannula, a speed of 1.2 ± 0.15 cm/s is observed near the nose, and a distance of 0.35 ± 0.02 m is reached before the exhalation jet loses linear momentum and become diffusive under shortness of breath conditions (*Q* = 4 Lpm). This is overall consistent with observations where nasal exhalation jets extending straight out to about 60 cm have been reported with adult humans that exhale somewhat greater *V*_*t*_ under normal breathing conditions [21].

When the nose and mouth are both open as in Fig. 1C, most of the exhalation emerges through the mouth, because of the relatively lower resistance offered by the wider and shorter oral passage compared with the nasal passage. The jet emerges from the mouth at a higher elevation angle of 36 ± 2 degrees in Fig. 1C compared with when it emerges from the nose as in Fig. 1B, under otherwise similar conditions. A greater speed of 1.64 ± 0.08 m/s, and greater distance of 0.51 ± 0.01 m is reached before the exhalation jet becomes diffusive when exhaling through the mouth compared with the nose. Because exhalations through the mouth are at a higher elevation angle, they reach a higher elevation compared to nasal breathing further increasing the risk to HCWs working near the patient.

To illustrate the dynamics, a backlit image of the exhalation jet emerging through the nose past the nasal cannula is shown in Fig 2A (from SI Movie 4), and the contained vortex dynamics made visible by the cross-sectional laser imaging in Fig. 2B (from SI Movie 5). Here the data corresponding to mid-range of flow rates used *Q* = 4 Lpm is shown. To illustrate the corresponding spread of the exhalation near the manikin, Fig. 2C shows the corresponding exhalation density averaged over several breath cycles projected in the vertical plane. The exhalation is observed to spread conically forward and concentrate in a single fast moving main jet as it mixes with the air in the room, loses momentum, and becomes diffusive. It can be noted that some secondary jets exist around the nose depending on exactly how the nasal cannula is mounted in the nose. Because of the presence of these jets, the exhalation density does not decay rapidly as inverse square of the distance from the nose/mouth of the patient, if it were spreading uniformly. Thus, the jets end up increasing the concentration of exhalations directly above and in front of the face of a supine patient.

**Figure 2:**
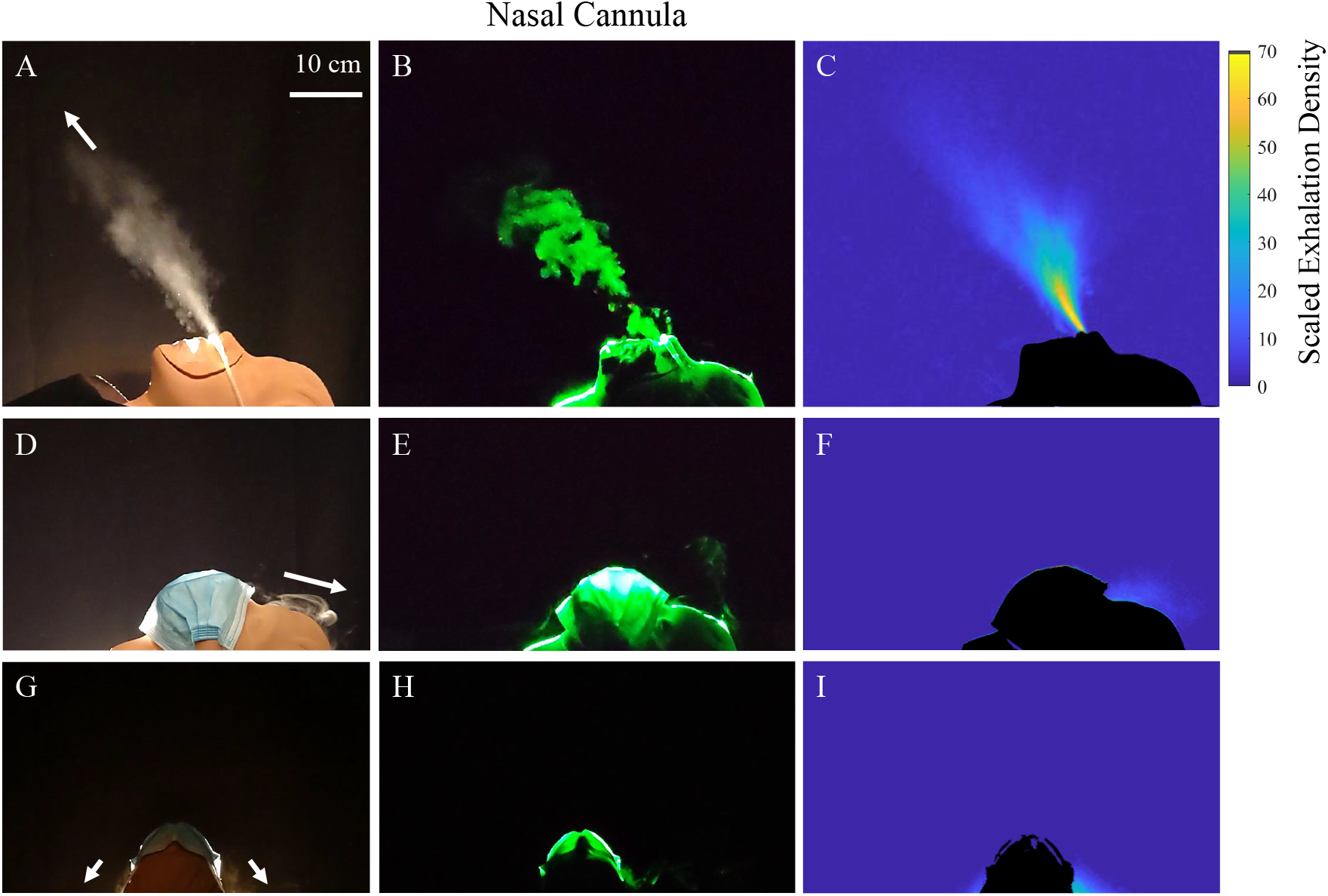
Exhalation jets emerge from the nose while wearing a nasal cannula with an oxygen delivery flow rate *Q* = 4 Lpm without a face mask (A-C). D-I: With a surgical mask on top, the exhalation jets are redirected mostly downward away from the face of a caregiver as indicated by the arrows (G-I), although some exhalations do escape from the bridge (D-F). The cross-sectional laser illumination (B) revels repeating pattern of swirling vortices signifying considerable linear momentum in the exhalation jets emerging past the unmitigated nasal cannula (see corresponding movie). The time-averaged projected exhalation density exhalation scaled by the mean density *ρ*_*m*_ = 3.49 10^*−*3^ kg/m^2^ if the exhalation were uniform is shown by the color bar.

### 3.2 Dispersal through Simple O2 Mask

Fig. 3A-C shows that a simple O2 mask redirects the exhalations largely through the vents on either side of the device, and to a smaller degree from the gap between the mask and the bridge of the nose. Here the data corresponding to mid-range of flow rates used *Q* = 8 Lpm is shown, and the corresponding Movies 6,7 show examples from Fig. 3A,B, respectively. Very little escapes from around the chin area because the O2 mask fits relatively tightly in that area. The exhalation jets from the vents on either side of the simple O2 mask appear broader and have a rounder shape compared with the jets coming from the nasal cannula. Vortex structures extending along lines starting at each of the vents are also evident from the cross section laser imaging on either side of the mask in Fig. 3B, and the associated Movie 7. Just as in the exhalation past the nasal cannula, these swirling vortices are associated with significant linear momentum when a fast-moving fluid enters a still region. The distance the jets extend out is approximately ± 0.29 0.02 m. These slightly lower distances compared to the nasal cannula are consistent with the formation of two dominant jets versus one dominant jet as in the nasal cannula.

**Figure 3:**
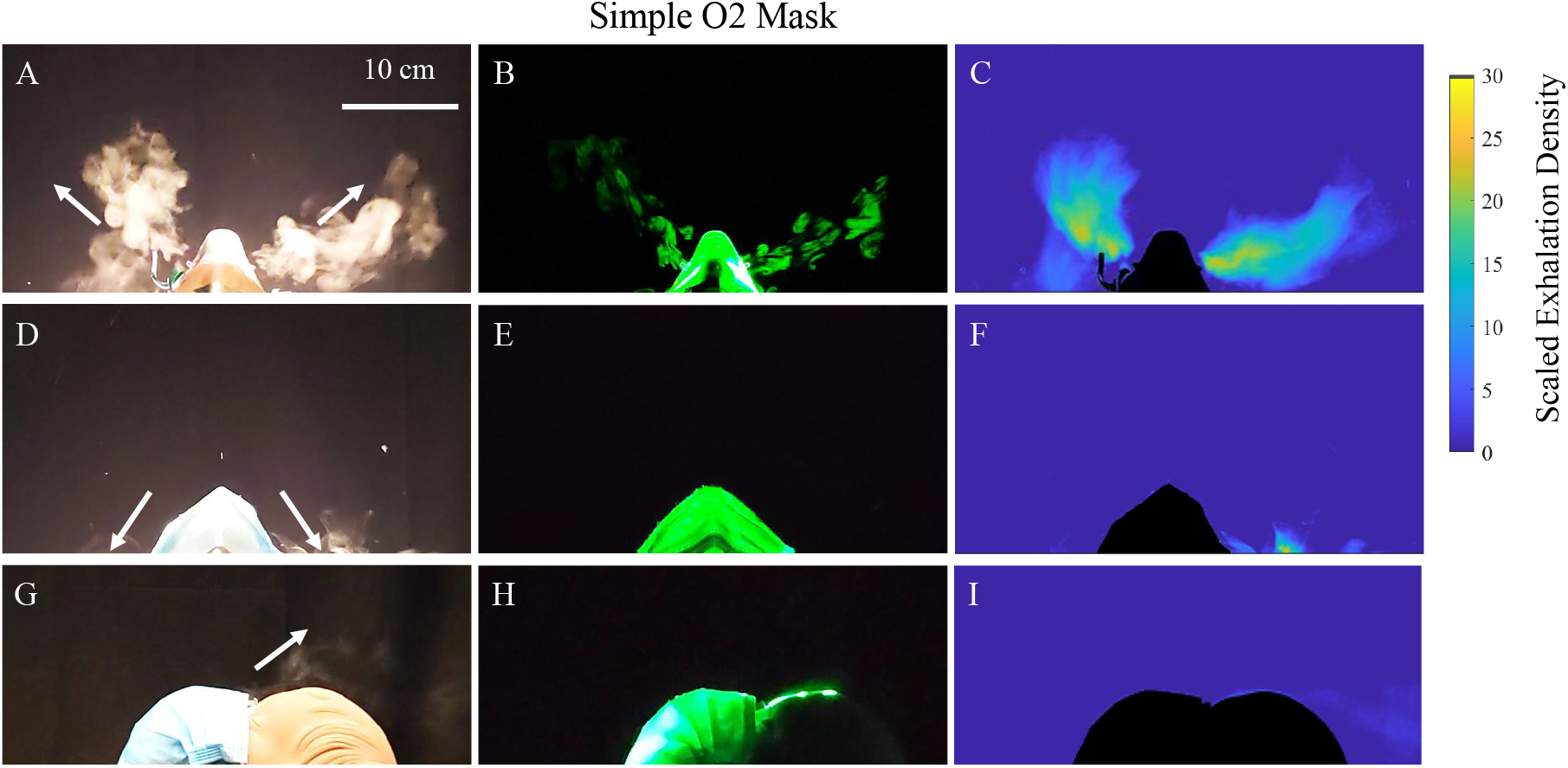
Exhalations emerging from the simple O2 mask during exhalation phase with an oxygen delivery flow rate *Q* = 8 Lpm without wearing a surgical mask (A-C), and while wearing a surgical mask (D-I). Note the repeating vortex structures along the jet emerging from the simple O2 mask vents indicates significant exhalation momentum (B). With a surgical mask on top, the jets are redirected mostly downward away from the face of a caregiver as indicated by the arrows, although some exhalations do escape from the bridge. The time-averaged projected exhalation density exhalation scaled by the mean density *ρ*_*m*_ = 1.41 10^*−*3^ kg/m^2^ if the exhalation were uniform is shown by the color bar.

The average exhalation density reached over an entire breathing cycle is shown in Fig. 3C, looking down from the top of the head. While not as elevated as in the case of the nasal cannula, the exhalation density resulting from the jets emerging from the vents in the simple O2 mask are pointed directly where HCWs typically stand while giving care to a supine patient. Because of the presence of the jets, the exhalation density in the case of the simple O2 mask does not decay as rapidly as the inverse of the square of the distance from the face, but rather is concentrated in particular directions. It can be further observed from the color bar that the exhalation densities reached near the vicinity of the head are comparable to those reached near the vicinity of the head while wearing a nasal cannula shown in Fig. 2C.

Thus, the exhalation density around an unmitigated oxygenation device is greater in different directions in relation to the head because of the presence of the jets in each device versus if the exhalations were diffusing out uniformly from the device. Hence, we examine the effect of placing a surgical mask to test its effect on exhalation dispersal.

### 3.3 Exhaled Jet Redirection with Surgical Mask Cover

Figure 3D-I shows the effect of placing a loosely fitted surgical face mask over an oxygenation device under otherwise similar conditions. Orthogonal views are shown to give a complete picture of the exhalation dispersal with this mitigation strategy. Here, the surgical mask was placed loosely on top of the oxygenation device to limit the effect on work of breathing. Thus, the primary objective is to deflect the exhalation jets, rather than to filter them as in the N95 mask. From these side-by-side images, and the associated movies in the supplementary documentation (SI Movies 6,7) over the range of oxygen flow rates used, we observe that the exhalation jets are reduced and deflected behind the manikin face.

Contrasting the unmasked and masked case in Fig. 2 and Fig. 3, the addition of the surgical mask atop the oxygenation devices works to dissipate the initial momentum of the exhaled air besides redirecting the exhalation jets coming through the devices downward. Thus, health workers who would otherwise be in the direct pathway of the exhalation when providing care to a supine patient, will not face the direct exhalation jet. Rather, the exhalation will be redirected downward to be nearly orthogonal to the directional line between a patient and their caregiver, significantly alleviating the direct exhalation concentration above the mask.

In order to quantify the degree of mitigation, Fig. 4A shows the angular exhalation density as a function of angle *θ* around the manikin head as defined in the inset to Fig. 4A. Here the angular density is obtained by integrating the measured projected exhalation density (as in Fig. 2C and Fig. 2F) from the face out to the furthest distance where exhalations are observed to reach above the face. Both plots corresponding to the exhalations without, and with, the surgical mask fixed atop the oxygenation devices are shown. It can be observed that the large jet which travels upward and forward is clearly suppressed by the placement of the mask. The same angular exhalation density is calculated for simple O2 mask using the views shown Fig. 3C and Fig. 3F, and plotted in Fig. 4B. Two peaks are observed corresponding to the two principal jets that emerge upward and outward from the vents of the simple O2 mask. As in the nasal cannula, clear suppression of the exhalation jets is quantitatively observed with the surgical mask placed over the simple O2 mask.

**Figure 4:**
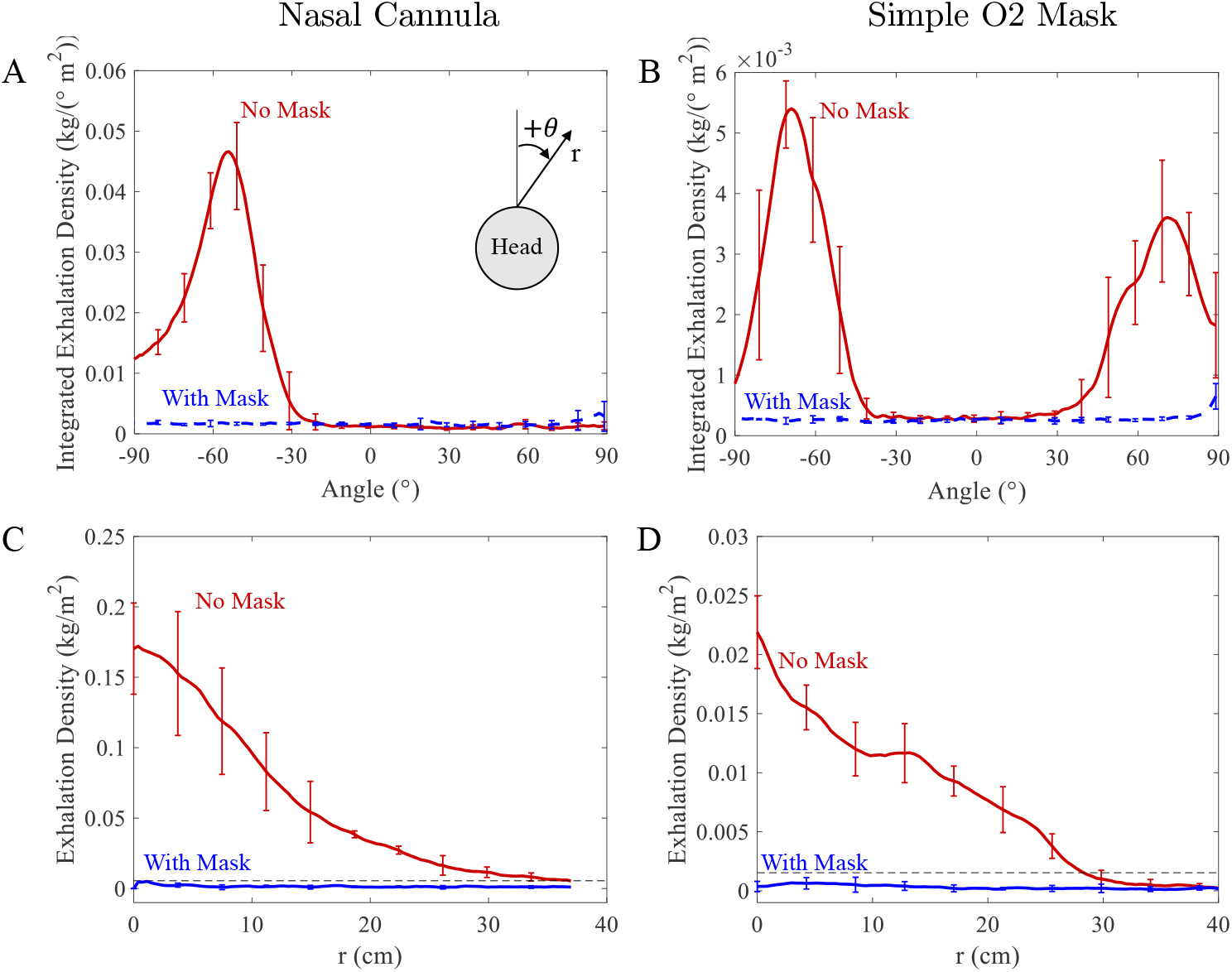
The angular integrated dispersal exhalation intensity is suppressed when a surgical mask is placed on top in both devices (A,B). The distance over which the jets reach is observed to be dramatically curtailed in both devices (C,D). The bars represent the range of values among five trials used to calculated the plotted averages. The exhalations travel up to 37 cm for the nasal cannula and 31 cm for the simple O2 mask when unmitigated. The black dashed line represents the threshold value that is 50% greater than the mean exhalation density *ρ*_*m*_ used to determine the exhalation distance.

To quantify the degree of mitigation with distance above the mask, we plot the exhalation density along the principal jets in the unmasked cases in the nasal cannula and simple O2 mask in Fig. 4C and Fig. 4D, respectively. These directions also correspond to the angle at which the maximum in the angular exhalation density occurs in each device in Fig. 4A and Fig. 4B. The exhalation density is observed to become significantly lower with the surgical mask on. Comparing the values without and with surgical mask in Fig. 4C,D, the exhalation density at *r* = 15 cm can be observed to be at least 30 times smaller in each device.

Thus, adding a surgical mask even loosely over either oxygen therapy devices can be seen to quantitatively reduce direct exposure to high exhalation aerosol concentrations created by exhalation jets above the face.

### 3.4 Dispersal mitigation with Oxygen flow rate

To quantify the degree of mitigation by a surgical mask with each oxygenation device, the percentage of exhalations observed above the face without and with a surgical mask is obtained by integrating the measured exhalation density above the plane defined by the surgical mask over one breath cycle. Figure 5A and Fig. 5B show the degree of mitigation observed over the oxygen flow rates examined in the nasal cannula and simple O2 mask, respectively, averaged over 5 independently measured breathing cycles in separate data runs. The percent of exhalations as a function of oxygen flow rates *Q* in Fig. 5 show no particular trend and can be considered more or less flat across the entire range even considering the small variations noted by the error bars from the five independent experiments. We further characterize the cone angle of the exhalation jet in both devices in Supplementary Information C, and show that it is essentially unchanged across flow rates and close to the universal value of 23.6°found for turbulent jet entry to a relatively still fluid and the law of similarity [26].

**Figure 5:**
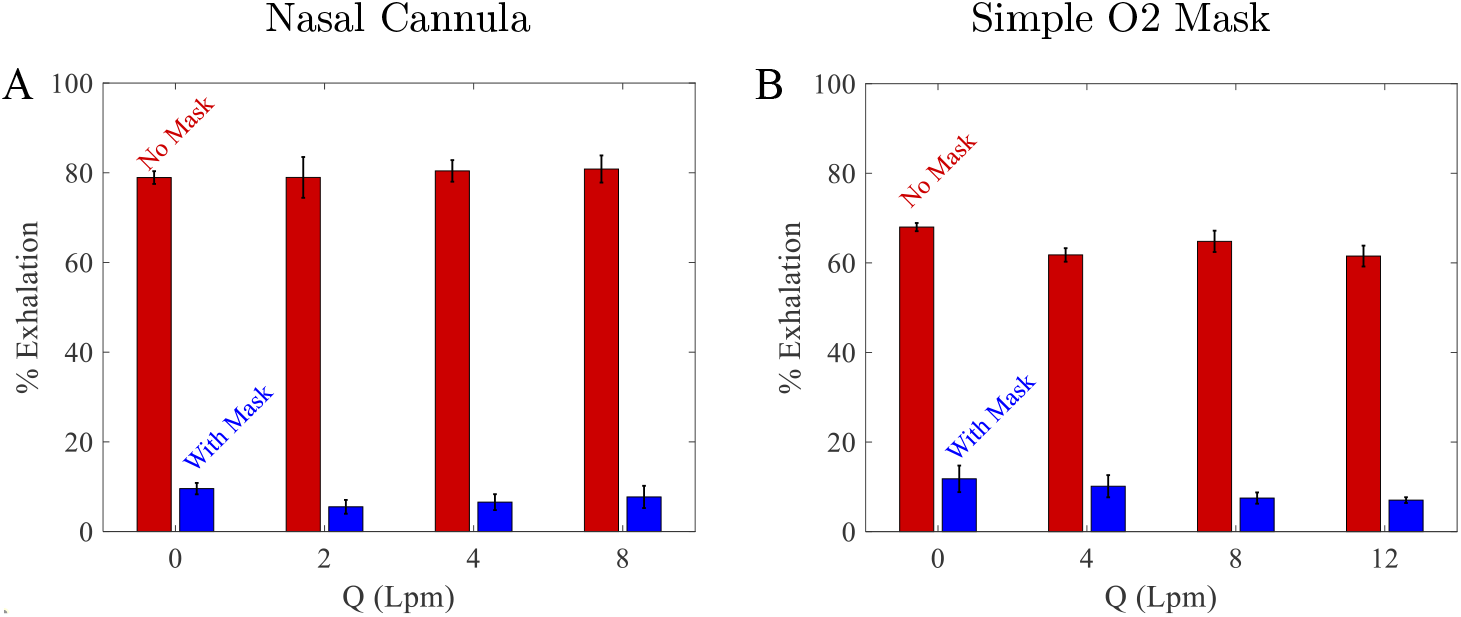
The percent of breath that is exhaled above the face without and with surgical mask as a function of flow rates for (A) the nasal cannula and (B) the simple O2 mask. All cases have five trials.

The average exhalation volume per minute given by the number of breaths per minute times the tidal volume, which is 7 Lpm under shortness of breath conditions, and 6 Lpm under normal breathing, are similar to *Q* in case of the nasal cannula. However, if one considers that the majority of the exhalation occurs over a fraction of the breathing cycle, the momentum associated with the exhalation itself can be proportionally higher and can dominate the dispersal over the range of oxygen flow rates used. Using the surgical mask limits the exhalations from the jets in the directions above the mask to about 10 % of the total exhalation volume over the entire range of oxygen flow rates. As noted in Fig. 2 and Fig. 3, 2nd and 3rd row, the exhalations do escape from the sides downward, and can diffusive up. However, this contribution to the exhalation with surgical mask is significantly less compared with unmitigated exhalations.

## 4 Discussion

Reproducible exhalations with a breathing simulator and manikin fitted with primary oxygenation devices clearly demonstrate fast moving jets that concentrate aerosol-laden exhalations directly above the nasal cannula and the simple O2 mask, where HCWs are typically positioned. While experiencing shortness of breath, aerosol-laden exhalations jet out mostly in the direction in which the manikin is facing through the nose and/or mouth. The strong exhalation jets result in significant aerosol exhalation concentrations above and beside a patient. This confirms that mitigation strategies are required to reduce the danger posed by the concentrated exhalation jets beyond wearing PPE. Mitigation strategies are required to further reduce the danger posed by the concentrated exhalation jets.

The simple O2 mask has three jets in total each in different directions: two large puff-like jets from the mask exhalation vents and one small jet outward from the bridge of the nose. The large jets from the vents on the side are directly where the face of a clinician would be positioned while giving direct care to the patient. The nasal cannula only has the one major jet coming directly straight out from the mouth, or nose. This makes it easier to avoid the jet of aerosols in front of the patient using a nasal cannula (when there is no mitigating surgical mask atop the oxygenation device,) because its path is more predictable. However, because these jets reach a greater height before turning diffusive, there is greater risk of further slow dispersal depending on the ventilation currents in the room. Mitigation in both cases is necessary to reduce risk of direct exposure of infectious aerosols to caregivers.

A loosely fitted surgical mask over a nasal cannula, or simple O2 mask, redirects exhalations downward and thus away from the faces of caregivers, while simultaneously reducing the volume and density of the exhalations reached above the patient. The surgical mask is only loosely placed to alleviate any concern for increased work of breathing. The surgical mask is demonstrated to quantitatively reduce exhalation density concentration above the mask. By preventing the exhalations from being launched directly up, the placement of a mask can also suppress wider dispersal of the exhalations depending on the ventilation currents. It should be emphasized that our study pertains to a supine patient, and does not apply to patients in the prone position.

In current practice, there is generally no mitigation in place on the patient side if they sneeze or cough while receiving care. The placement of the mask can also reduce the larger aerosol and droplets expired if the patient speaks, coughs or sneezes if not totally eliminate them [20, 21, 28, 23].

Our main findings are summarized as follows:

- When using the nasal cannula the exhalation jets to 0.35 ± 0.02 m, and split slightly around the stem of the device while nasal breathing. The jet is angled slightly upwards in comparison to the free breathing case, and its angle varies somewhat depending on exactly how the nasal cannula is placed. Mouth breathing angles the exhalations are relatively higher compared with nasal breathing with a nasal cannula, leading to greater dispersal distances.
- The simple O2 mask has upward and lateral jets that travel out to 0.29± 0.02 m from the mask vents on both sides, and one smaller upward jet from the bridge of the nose.
- The simple O2 mask has three jets whereas the nasal cannula has one jet making it easier to stay out of the way of the initial momentum of the jet for the nasal cannula. However, the exhalations are launched higher and thus spread further as they slowly disperse depending on the air flow within the room.
- The surgical mask reduces and redirects the jets for the nasal cannula and the simple O2 mask while dissipating the momentum of the exhalations. The surgical mask is found to limit the direct exhalations to about 10 % above the mask over the entire range of oxygen flow rates used in either device.

## 5 Conclusions

We demonstrate that significant exhalation jets exist with either a nasal cannula or simple O2 mask commonly used in treating COVID-19 patients, giving rise to high exhalation concentration above and around a supine patient, with quantitative measurements enabled by a manikin breathing simulator. Mitigation is demonstrated by reducing and redirecting exhalations by using a surgical mask over the oxygenation devices. In all cases the exhalation jets are directed downwards and away from the faces of HCWs working around the patient’s head. The build up of exhalation density due to jets are found to be alleviated by nearly 90% with a surgical mask placed above both oxygenation devices.

This study demonstrates the efficacy of placing a simple surgical face mask over a supplemental oxygen delivery device (Nasal Cannula/Simple O2 Mask) in reducing aerosol exposure risk to healthcare workers treating patients with COVID-19, and other infectious respiratory diseases.

## Data Availability

All data is contained in the manuscript.

http://physics.clarku.edu/~akudrolli/covid

## Acknowledgments

We thank Chris Blanker, Germano Iannacchione, and Catherine Taylor for advice, discussions, and help with study. This work was supported by U.S. National Science Foundation COVID-19 RAPID Grant No. DMR-2030307.

