## Supplementary Information for "Assessment & mitigation of O2 therapy driven spread of COVID-19"

### Supplementary Information
### Assessment & mitigation of O2 therapy driven spread of COVID-19

February 6, 2021

Arshad Kudrolli[a], PhD, Brian Chang[a], PhD, Jade Consalvi[a], Anton Deti[a], Christopher Frechette[b], RRT, Helen Scoville[b], RRT, MSM, Geoffrey R. Sheinfeld[c], MD, and William T. McGee[b], MD, MHA
[a] Department of Physics, Clark University, Worcester, MA 01610; [b] University of Massachusetts Medical School-Baystate, Springfield, MA 01107; [c] Andover, MA 01810.

#### A Oxygenation devices

We examine two commonly used oxygenation devices: the simple $O_2$ mask and the nasal cannula used in respiratory treatment Pyramid shown in Fig. S1. The simple $O_2$ mask covers both the nose and mouth of the patient, but has two vents that would allow air to freely pass in and out of the mask. Meanwhile, a steady oxygen supply is delivered through the mask at a rate of $Q$ ranging between 4 Lpm and 12 Lpm. The nasal cannula is a tube that splits into two prongs that are partially inserted into the nose. Oxygen is then delivered through the nose at flow rate $Q$ ranging between 2 Lpm and 8 Lpm.

#### B Image Analysis

We use two complementary visualization methods to obtain an integrated view of the direction and spread of the exhalation over several breathing cycles building off methods which we and other groups have used previously in visualizing exhalations. In previous work, we demonstrated that a standard nose and mouth mask reduces the mucosaliva dispersed by a factor of at least a hundred compared to the peaks recorded when unmasked [1]. In this study we use further laser illumination similar to those used previously [2, 3] and a lung simulator from Michigan Medical Systems with breathing volume and periods corresponding to normal breathing, and those with shortness of breath. An aerosol-laden exhalation jet emerging from the nose of the manikin while free breathing imaged with laser illumination is shown in Fig. S2B.

The videos are analyzed using the MATLAB image processing toolbox. The mean measured light intensity corresponding the light scattered by the aerosol-laden exhalations after a exhalation cycle is mapped to the mean projected exhalation density $\rho_m$ in the measured frame encompassing the entire area over which exhalations are observed to reach in one breathing cycle. This mean density is given by the mass of the exhaled volume of air $V_t$ multiplied by the density of exhaled air $\rho_a = 1.22$ kg/m$^3$, and divided the area of the frame. This density corresponds to the density if the exhalation were uniform spread, and can be used to access the relative risk of higher dose of virus bearing exhalations in a certain area because of the jets compared if the exhalations spread out uniformly. The distance where the exhalation jets reach is identified by plotting the scaled exhalation density along the observed jets, and then identifying the point where the density has decreased to be within 50% of the mean density.

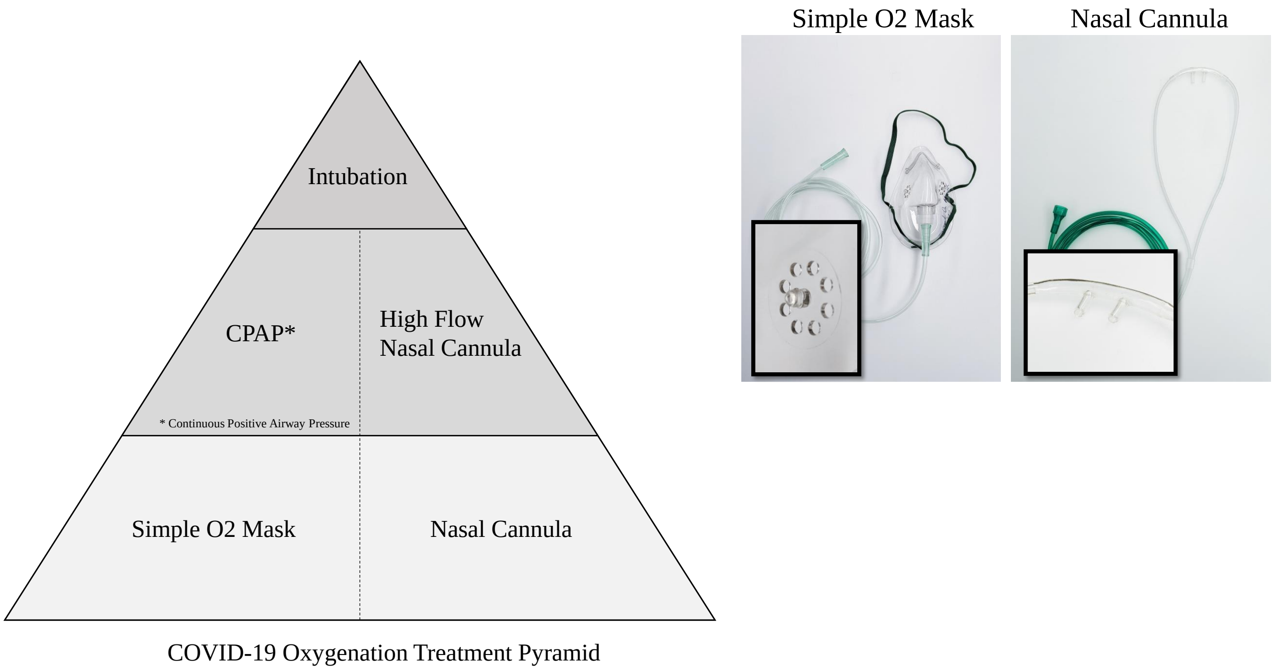


Figure S1: The simple O2 mask and nasal cannula are used interchangeably in terms of oxygenation efficacy at the lower level of the respiratory disease treatment pyramid. Images of the oxygenation devices used in the study with insets showing closeups of the nozzles in each device.

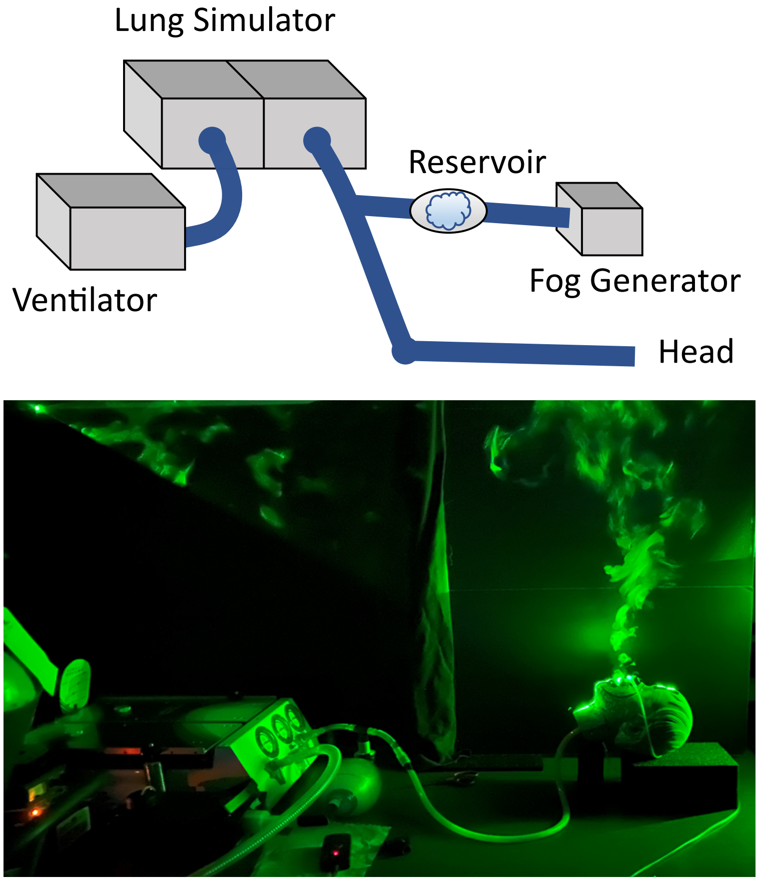


Figure S2: The study system consists of a dual lung simulator driven by a ventilator coupled with a second lung chamber which “breathes" air through the manikin head shown with a prescribed frequency and tidal volume. The exhalations are visualized using water-glycerol aerosols (fog) released through the manikin mouth and/or nose during exhalation. Image of system with laser-sheet lighting used for aerosol visualization.

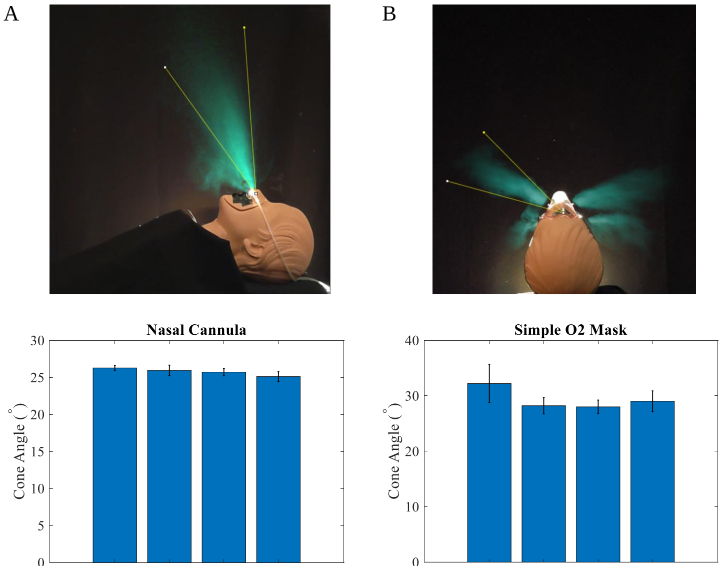


Figure S3: Representative time averages showing the cone angles of the exhalations jets for the nasal cannula (A) and simple O2 mask (B). The measured exhalation angles are observed to be essentially constant across the oxygenation flow rates. The data for $Q = 0\,\mathrm{Lpm}$ was taken for reference.

### C Breath Exhalation Physics

When an exhalation jet from the mouth or nose enters the relatively quiescent air it spreads out in a cone in the time-averaged images as shown Fig. S3. Turbulent exhalation jets emerging from the nose or mouth during breathing are called puffs by fluid mechanics researchers when they emerge periodically or as a pulse [4], whereas a jet refers to a continuous stream of fluid at high Reynolds number. The Reynolds number is used to characterize the physics nature of fluid flow is given by $\mathrm{Re} = \frac{DU}{\nu}$, where $\nu$ is the kinematic viscosity which is $1.6 \times 10^{-5}$ m$^2$/s for air, $D$ is the jet diameter, and $U$ is its speed. Assuming, $D \approx 2\,\mathrm{cm}$, and $U \approx 1.5$ m/s, $\mathrm{Re} \approx 2000$ [5]. The flows corresponding to these Re are considered turbulent, consistent with the observation of vortex swirl patterns seen in Figures 2 and 3, and related movies as the jet enters the relatively still air. It must be noted here that all the repeated structures is not simply a result of periodic breathing, but the result of vortex shedding [6].

The measured cone angle of the turbulent exhalation jets are shown in Fig. S3. The measured angles are observed to be nearly constant across the oxygenation flow rates showing that the crosssection of the jets are not affected by the oxygen flowing from the nasal cannula in the nose. This is also consistent with the near constant percentage of exhalation observed to move upward and forward as a function of flow rates in the nasal cannula. When a fast moving jet enters a still fluid from a uniform conduit, the universal value of $23.6^o$ based on the law of similarity [6]. The cone angles do not change with oxygenation rate in neither the nasal cannula nor the simple O2 mask in Fig. S3. The cone angles were measured to be close to the universal value for $26.4 \pm 1.5$ for nasal breathing, and $26.6 \pm 1.5$ for mouth breathing considering the error in measurements. Thus, we deduce that the flow around its stem appears to lead to a slightly larger cone angle compared to the universal value.

**Acknowledgments** We thank Chris Blanker, Germano Iannacchione, and Catherine Taylor for advice, discussions, and help with study. This work was supported by U.S. National Science Foundation COVID-19 RAPID Grant No. DMR-2030307.

[6] Benoit Cushman-Roisin and Jean-Marie Beckers. *Introduction to Geophysical Fluid Dynamics - Physical and Numerical Aspects*. Academic Press, 2005.

**List of Movies**

- Movie 1: Mannikin supine simulating shortness of breath through the nose ($V_t = 350$ mL and $f = 20$ bpm) with a nasal cannula at $Q = 4$ Lpm.
- Movie 2: Mannikin supine at 45° simulating shortness of breath through the nose ($V_t = 350$ mL and $f = 20$ bpm) with a nasal cannula at $Q = 4$ Lpm.
- Movie 3: Mannikin supine simulating regular breathing through the mouth ($V_t = 500$ mL and $f = 12$ bpm) with a nasal cannula at $Q = 4$ Lpm.
- Movie 4: Movie corresponding to Fig. 2A,D.
- Movie 5: Movie corresponding to Fig. 2B,E.
- Movie 6: Movie corresponding to Fig. 3A,D.
- Movie 7: Movie corresponding to Fig. 3B,E.
